# Machine learning based phenotyping of the response to mindfulness for chronic low back pain

**DOI:** 10.1101/2025.08.19.25333990

**Authors:** Akshit Soota, Angela C. Incollingo Rodriguez, Benjamin C. Nephew, Paula Gardiner, Jean A. King, Natalia Morone, Carolina Ruiz

**Affiliations:** Worcester Polytechnic Institute, Worcester, MA; Cambridge Health Alliance, Cambridge, MA; Boston Medical Center, Boston, MA

## Abstract

Millions of people each year suffer from chronic low back pain (cLBP), which adversely affects their physical and mental health. While non-pharmacological interventions such as mindfulness are known to be effective in treating cLBP, not all patients experience the same benefit. Determining who these treatments might work best for is difficult, as there are no reliable predictors of the response to mindfulness for cLBP. The objective of the current study was to apply predictive machine learning to data collected from a completed clinical trial of mindfulness for cLBP to identify phenotypes characterizing those who did and did not respond to the intervention. The analyses here focused on 132 participants in the intervention arm of the clinical trial of mindfulness for cLBP. The Random Forest machine learning technique was used to identify key characteristics of responders (49) and non-responders (83). The top three responder phenotypes were able to identify 26 out of the 49 responders with 92-100% precision. The top three non-responder phenotypes were able to identify 36 out of 83 non-responders, all with 100% precision. Results from this machine learning based phenotyping can guide clinician and patient decision-making to maximize clinical efficiency, patient outcomes, and resource use as well as inform research and development of mindfulness-based treatments for pain.

## INTRODUCTION

Chronic low back pain (cLBP), defined as pain lasting three months or longer, affects approximately 9–10 % of adults worldwide, with prevalence increasing from early into late adulthood (Meucci et al., 2015). Globally, over half a billion people reported LBP in 2020, and chronic cases are expected to increase by more than 36 %—to over 843 million—by 2050 as populations age globally (Ferreira et al., 2023). As the leading cause of disability worldwide, cLBP is a substantial cause of lost productivity and imposes significant economic burdens, with individual costs reaching $10-20,000 per patient per year (Fullen et al., 2022). Beyond the financial toll—estimated at hundreds of billions annually in many countries—affected individuals frequently experience reduced quality of life, higher rates of depression, early retirement, and increased mortality risk (The Lancet, 2023).

Standard treatment protocols for cLBP typically begin with nonpharmacologic interventions, which are favored due to their lower risk profiles and modest but consistent benefits. The American College of Physicians and related guidelines strongly recommend exercise programs (e.g., aerobic activity, motor-control/core strengthening), mind body interventions (mindfulness-based stress reduction, yoga, Tai chi), and multidisciplinary rehabilitation, each supported by moderate-quality evidence demonstrating small to moderate improvements in pain and function (Hauk, 2017). If these nonpharmacologic methods are not effective, guidelines suggest progressing to oral NSAIDs as first-line medications, which are moderately effective for many patients. Second-line pharmacologic agents include SNRIs (notably duloxetine), tramadol, and tricyclic antidepressants, each offering small improvements in pain relief (Korownyk et al., 2022). However, while opioids may provide short-term pain reduction, they are not always advantageous. These medications carry considerable risk profile—withdrawals, adverse effects, and potential for misuse— and are thus reserved for cases where other therapies have failed and after careful consideration by patient and provider. The ongoing opioid epidemic (Volkow and Blanco, 2021), with over 54,000 opioid related overdose deaths in 2024 alone (Post et al., 2025), has increased interest and efforts to maximize the application and effectiveness of nonpharmacologic interventions, including mindfulness.

Mindfulness-based interventions, particularly Mindfulness-Based Stress Reduction (MBSR) and related meditation practice, are often effective, low-risk treatments for cLBP. A comprehensive meta-analysis of 12 randomized controlled trials involving 1,153 patients found that meditation-based therapies modestly but significantly reduced pain intensity and bothersomeness while improving quality of life compared with non-meditation controls (Lin et al., 2022). Additional systematic reviews of MBSR have reported short-term reductions in pain intensity (mean difference ≈ 1 on a 0–10 scale) and modest gains in physical function, though long-term superiority to other active treatments is unclear (Anheyer et al., 2017). A large randomized trial (n=342) comparing MBSR, Cognitive Behavioral Therapy (CBT), and usual care found that both MBSR and CBT yielded greater improvements in pain and functional limitations at 26 and 52 weeks—with MBSR effects persisting at one year—versus usual care (Cherkin et al., 2016, Cherkin et al., 2017). Overall, mindfulness programs for cLBP consistently yield small to moderate benefits in pain, function, and well-being, underscoring their role as a valuable component of nonpharmacologic, multimodal pain management strategies.

Machine learning (ML) techniques can be used to effectively predict and evaluate pain outcomes across a spectrum of clinical scenarios. A 2021 review of 26 predictive ML studies of pain outcomes (diagnosis, pain intensity, managing pain) reported superior performance compared to traditional statistical methods (Matsangidou et al., 2021). Similarly, a systematic review of 44 EEG-based ML studies reported that models could accurately predict pain intensity and treatment response (65-100% accuracy) demonstrating substantial analytic potential depending on the method used (Mari et al., 2022). One 2024 study using high-dimensional clinical data from over 67,000 outpatient records found that ensemble ML methods such as Random Forest outperformed standard logistic regression and other machine learning methods in predicting chronic pain presence and treatment response—highlighting key predictors such as BMI and inflammatory markers (Wu et al., 2024). In summary, there is consistent evidence of ML’s effectiveness in predicting pain intensity and treatment outcomes, but it has not been applied to predict the response to mindfulness-based interventions for pain.

The objective of the current secondary data analysis study was to apply predictive ML to data collected from a completed randomized clinical trial of mindfulness for cLBP (the Aging Successfully with Pain study)(Morone et al., 2016). This parent clinical trial sought to determine the effectiveness of a mind-body program at increasing function and reducing pain in older adults with cLBP. The present analyses extended this work to identify clusters of participant features (hereafter termed “phenotypes”) characterizing those who did and those who did not respond to the intervention. Analyses focused on the intervention arm of the RCT and used Random Forest to identify phenotypes of responders and non-responders.

## METHODS

### Aging Successfully with Pain RCT Overview

This experimental study was designed as a randomized, patient education-controlled clinical trial of a mind-body program for older adults with cLBP. Details of the study procedure have been published previously (Morone et al., 2016, Morone et al., 2012). 282 independent, community-dwelling adults 65 years or older were recruited from metropolitan Pittsburgh, Pennsylvania. Participants in the mind-body (intervention) group received the group intervention of 8 weekly 90-minute mindfulness meditation sessions modeled on the Mindfulness-Based Stress Reduction program (Kabat-Zinn, 2003, Kabat-Zinn and Hanh, 2009). Controls received an 8-week group health education program based on the “10 Keys” to Healthy Aging. After completion of the 8-week program, participants in the intervention and control programs were asked to return for 6 monthly booster sessions. Measures were obtained at baseline, after the 8-week program, and 6 months after program completion.

Recruitment occurred from February 14, 2011, to June 30, 2014. The final 6-month assessment was completed April 9, 2015. The study protocol was approved by the institutional review board of the University of Pittsburgh. All participants provided written informed consent.

### RCT Inclusion and Exclusion Criteria

Participants were included if they were 65 years or older, spoke English, had intact cognition (Mini-Mental State Examination score, ≥24)(Folstein et al., 1975), had functional limitations owing to their chronic LBP (defined as a score of ≥11 on the Roland and Morris Disability Questionnaire [RMDQ] (Roland and Morris, 1983); range, 0-24, with higher scores indicating increased limitations), and had self-reported moderate chronic pain levels on a verbal descriptor scale (Pain Thermometer; measured on a visual scale as pain as bad as it could be, extreme, severe, moderate, mild, or no pain) occurring daily or almost every day for at least the previous 3 months (Roland and Morris, 1983). Participants were excluded if they had participated in a previous mindfulness meditation program, had serious underlying illness (such as malignant neoplasms, infection, unexplained fever, weight loss, or recent trauma) causing their pain, were nonambulatory, had severe impaired mobility, had visual or hearing impairment that interfered with assessments, had pain in other parts of the body more severe than their chronic LBP or acute back pain, had an acute or a terminal illness, or had moderate to severe depressive symptoms (Geriatric Depression Scale score, ≥21; range, 0-30) (Yesavage et al., 1982).

### RCT Assessments and Outcome Measures

The Roland-Morris Disability Questionnaire (**RMDQ**) was the primary outcome measure. It contains 24 questions specifically related to functional limitations as a result of LBP. A clinically meaningful change in the RMDQ ranges from a 2.5– to a 5.0-point improvement (reduction) from baseline (Kovacs et al., 2007).

Pain (present, average, and most severe during the past week) was measured by self-report with the 21 point Pain Numeric Rating Scale (**PNRS**; range, 0-20, with higher scores indicating worse pain)(Herr et al., 2004). Because pain is a complex phenomenon that affects quality of life, mood, and psychological function, a variety of established instruments were used to measure these different domains.

The Modifiable Activity Questionnaire (**MAQ**) was used to record the frequency and duration of various levels of physical activity (Kriska et al., 1990). The MAQ assesses activities during occupational and leisure time, as well as inactivity due to disability. A total score is calculated by summing the scores from each domain, ranging from 3 to 15, where a higher score indicates a higher level of physical activity.

Quality of life was measured with the RAND-36 Health Status Inventory (**SF-36**, consisting of the physical functioning, mental, health, and general health perception scores, with higher scores indicating better health (Hays and Morales, 2001).

The Profile of Mood States (**POMS**) was used to measure transient and fluctuating feelings and enduring affect states. It’s a 65 item self-report questionnaire that evaluates six mood states: tension-anxiety, depression, anger-hostility, vigor, fatigue, and confusion (McNair et al., 1971).

Given the strong association between chronic pain and depression, the 30 item Geriatric Depression Scale (**GDS**) was used to assess depression (Magni et al., 1990). It has a score range of 0-15, with higher scores indicating more severe depression. Scores are typically interpreted as follows: 0-4 = normal, 5-8 = mild depression, 9-11 = moderate depression, and 12-15 = severe depression. Self-efficacy has been shown to predict task performance (Bandura, 1984). This construct was measured with the well-validated Chronic Pain Self-Efficacy Scale (CPSE, range, 0%-100%, with higher scores indicating improved self-efficacy)(Anderson et al., 1995).

The Fear Avoidance Beliefs Questionnaire (**FABQ**) was used to assess how fear-avoidance beliefs about physical activity and work affect and contribute to low back pain (Waddell et al., 1993). The FABQ consists of 16 items in which an individual rates their agreement with each statement on a 7-point Likert scale. There is a maximum score of 66, where a higher score indicates more strongly held fear avoidance beliefs.

Chronic pain acceptance was measured with the Chronic Pain Acceptance Questionnaire (**CPAQ**)(McCracken et al., 2004), which assesses how individuals accept their chronic pain and its impact on their lives. The standard CPAQ has a range of 0-120, with higher scores indicating greater acceptance. The CPAQ has two subscales: Activity Engagement (score range 0-66) and Pain Willingness (score range 0-54). Each item is rated on a 7-point scale from 0 (never true) to 6 (always true). Pain catastrophizing was measured with the Catastrophizing Scale of the Coping Strategies Questionnaire (**CSCS**, range, 0-6, with higher scores indicating greater catastrophizing) (Rosenstiel and Keefe, 1983).

Self-reported mindfulness was assessed with the Mindful Attention Awareness Scale (**MAAS**; range, 1-6, with higher scores indicating greater mindfulness)(Brown and Ryan, 2003). Data on comorbidity were reported with the Cumulative Illness Rating Scale (range, 0-13, with higher scores indicating more comorbid conditions)(Linn et al., 1968).

The Multidimensional Scale of Perceived Social Support (**MSPSS**) is a 12 question measure used to measure perception of support from 3 sources: family, friends and a significant other (Zimet et al., 1988). The total score ranges from 12 to 84, with higher scores indicating greater perceived social support. Each subscale (family, friends, and significant others) also has a score range of 4 to 28.

The Credibility Expectancy Questionnaire (**CEQ**) is a six-item scale which was used to assess participants’ perceptions of treatment credibility and their expectancy of its effectiveness, with higher scores indicating greater credibility and stronger outcome expectations (Devilly and Borkovec, 2000).

### Objective measure

The Short Physical Performance Battery (**PPB**) (Guralnik et al., 2000, Guralnik et al., 1995, Guralnik et al., 1994) tests lower extremity function by measuring standing balance, gait speed and timed chair rise, tasks that are commonly encountered by older adults. The maximum score is 12 and higher scores indicate better function.

### Intervention

The intervention was modeled on the 8-week Mindfulness-Based Stress Reduction (MBSR) program (Kabat-Zinn and Hanh, 2009, Kabat-Zinn, 2003). Four methods of mindfulness meditation were taught: the body scan, sitting practice, walking meditation, and mindful stretching. These techniques take regular activities such as sitting, walking, and lying down and transform them into a meditation through directed breathing and mindful awareness of thoughts and sensations. To encourage proficiency with the meditation method after completion of the intervention, monthly 60-minute booster sessions were held. Each session included time for a mindfulness meditation and time for discussion of the themes brought up during the 8-week program.

## MACHINE LEARNING MODELING: DATASET

The final sample size for the RCT was 282 participants, with 140 participants in the intervention group and 142 participants in the control group. The intervention group was 34% male, had a mean age of 75, and was 70% white and 30% black. Our analyses focus on **132** out of 140 intervention participants with data at baseline and 8-weeks. The main outcome was the Roland-Morris Disability Questionnaire (RMDQ), which measures functional limitations owing to low back pain. Other self-reported questionnaires were collected by the study at baseline, 8-weeks (end of treatment), 6-months, and 12-months. Individual items from questionnaires, subscores and total scores were all used for the ML analyses.

## OUTCOME ATTRIBUTE: CLINICALLY SIGNIFICANT CHANGE IN RMDQ

The machine learning modeling results in the present analyses focus on the Roland-

Morris Score (RMscore). We used change in the Roland-Morris score as our predictive outcome, which we denote as ΔRMscore (“delta RMscore”) and compute by subtracting a subject’s 8-week score from their baseline score: Δrmscore = RMscore_baseline_ – RMscore_8-week_.

A positive ΔRMscore indicates that the subject saw an improvement in the self-reported disability measure. A within-patient change of at least 4 or 5 points in the Roland-Morris score is recognized as the threshold for a clinically important. Hence, in the current study our main outcome is a *binary treatment response outcome*, where ΔRMscore > 4 is considered clinically significant improvement (outcome class 1) and ΔRMscore ≤ 4 is *<u>not</u>* considered clinically significant improvement (outcome class 0). The number of participants with ΔRMscore > 4 is 49; and with ΔRMscore ≤ 4 is 83.

## MACHINE LEARNING MODELING TECHNIQUES

The modeling goal was to identify markers that predict clinically significant response (or lack of it) to treatment. The current analyses present markers and combinations of markers that our Random Forest based ML modeling has identified as predictive of treatment response. The *Random Forest (RF)* method constructs a collection of decision trees by including some randomness in the tree construction process that allows the creation of different decision trees over the same dataset; this reduces overfitting (Rokach, 2010). The Decision Tree algorithm is an ML method of choice for its high accuracy, descriptiveness, and fast training and execution time. Given a dataset, the algorithm builds a decision tree, which is a hierarchical representation of the predictive data variables in the order in which they should be tested to maximize predictive performance.

The algorithm determines the most predictive data variable (take for example “gait speed”) together with an optimal threshold to split the variable values (take for example “0.6”) using an optimization entropy-like metric (e.g., gini). The algorithm then creates a root node that contains this most predictive variable (gait speed) and creates two children of the node: the left child is for participants whose variable value is less than or equal to the threshold (gait speed ≤ 0.6) and the right child corresponds to the remaining participants (gait speed > 0.6). Each branch of the tree continues to be expanded in a similar way: the next node on the leftmost branch is the variable that is the best outcome predictor (take for example, fatigue) for participants on that branch (participants with gait speed ≤ 0.6) according to the optimization metric. The expansion of a branch terminates on a “leaf” node when there is sufficient certainty (as measured by a confidence metric) to make a prediction for participants who satisfy all variable “tests” on the branch. Here, that prediction (“class”) is a yes/no answer to whether a cLBP participant who matches all variable tests on the branch will respond to MBSR.

## PREDICTIVE PATTERNS: “PHENOTYPES”

We extracted the most predictive tree branches from the Random Forest obtained by our ML modeling. Each of these tree branches consists of a combination of baseline markers (such as participant features or study variables) that is highly predictive of response to treatment at 8-weeks. We call these patterns *“phenotypes”*. Each of these phenotypes can be represented as an IF-THEN rule of the following type:

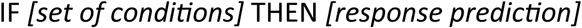

where “*set of conditions”* consists of one or more conditions on participant features (e.g., `Gait Speed > 0.6`); and the response prediction is either “yes” (i.e., clinically significant response to treatment) or “no” (i.e., no clinically significant response to treatment). The order in which these conditions appear in the phenotype corresponds to the predictive importance of the conditions, with the first condition being the most important in the prediction.

## METRICS USED TO MEASURE PREDICTIVE PERFORMANCE: PRECISION AND SENSITIVITY

Given that each IF-THEN phenotype makes only one type of prediction, either “yes” (i.e., clinically significant response) or “no” (i.e., not clinically significant response) for all the participants that satisfy the set of conditions in the phenotype. For example, the fake rule:

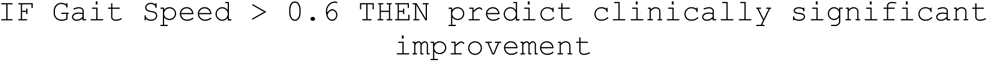

predicts clinically significant improvement for all participants whose gait speed is greater than 0.6, but it remains silent about the outcome of participants with gait speed ≤ 0.6. Hence, metrics such as accuracy, AUC, sensitivity, and specificity are not appropriate to measure the predictive performance of an IF-THEN phenotype. We consider instead two performance metrics, precision and recall, that best describe the predictive performance of these phenotypes:

- ***Precision:*** *Percentage of correct predictions among all predictions made by the phenotype.* In our case, this is the percentage of participants with the same outcome as the THEN-part of phenotype among participants who satisfy the condition(s) on the if-part of the phenotype.
- ***Sensitivity*** *(also called “Recall”): Percentage of correct predictions made by the phenotype among those whose outcome is the same as that on the THEN-part of the phenotype.* In our case, this is the percentage of participants who satisfy the conditions on the IF-part of the phenotype among all participants with the same outcome as the THEN-part of phenotype.

*Precision* is our main metric to measure the predictive performance of single phenotypes.

## RESULTS

### MARKERS PREDICTIVE OF RESPONSE: FEATURE IMPORTANCE

This section presents individual features that were deemed important by the Random Forest models in the prediction of the binarized intervention response. Figure 1 depicts the top 10 features identified by Random Forests as significant for predicting the RMDQ response to the mindfulness intervention. Each of these features appears frequently in the decision trees of the Random Forest model. Each of these features is not a good predictor of outcome by itself but is instead a significant contributor to combinations of features that together predict outcome. The following sections focus on analyzing the tree branches in the Random Forest to identify combinations of features that together have high predictive power.

**Figure 1:**
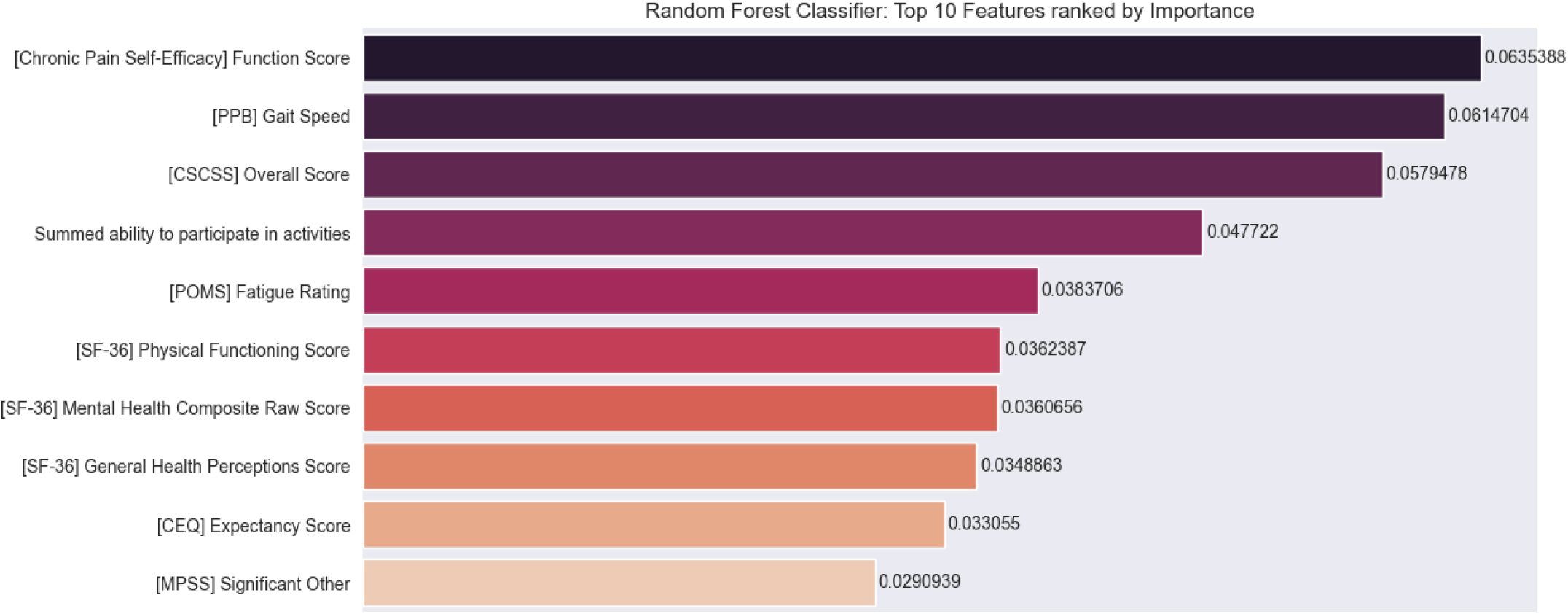
Top 10 features for predicting binary response to intervention. Chronic Pain Self-Efficacy (CPSE) function score; PPB (Short Physical Performance Battery) gait speed score; CSCSS (Catastrophizing Scale of the Cognitive Strategies Questionnaire) overall score; summed ability to participate in activities; POMS (Profile of Mood States) fatigue rating score; SF 36 (Short-Form Health Survey) physical functioning score; mental health composite score, and general health perception score; CEQ (Credibility Expectancy Questionnaire) expectancy score; and MPSS (Multidimensional Scale of Perceived Social Support) significant other score.

### COMBINATIONS OF MARKERS PREDICTIVE OF RESPONSE: PHENOTYPES

This section presents the top ranking IF-THEN phenotypes identified with Random Forests to predict whether the change in Roland-Morris score from baseline to 8-weeks was a clinically significant improvement (i.e., Δrmscore > 4 for responders) or not (i.e., Δrmscore ≤ 4 for non-responders). A random sample of participants and a random sample of input features were used to construct each tree in the forest. The number of participants who were included in the construction of the decision tree that yielded each of the phenotypes is reported below.

#### Top 3 responder phenotypes

##### Responder phenotype 1 improvement in Roland Morris with MBSR

**Figure 2:**
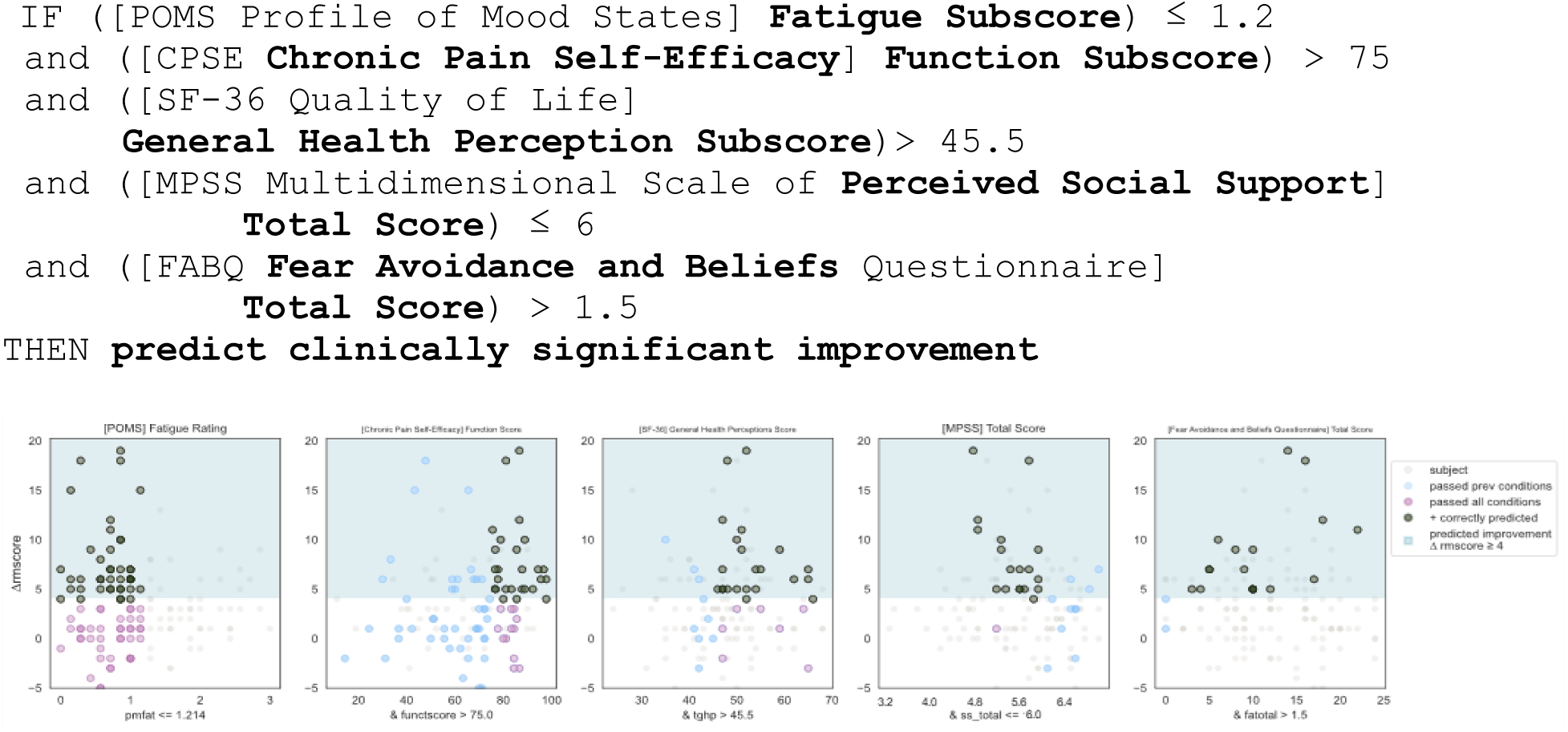
Responder Phenotype 1 Scatter Plots. Each scatter plot corresponds to one phenotype condition and depicts the phenotype condition values on the x-axis vs. the outcome (ΔRMscore) values on the y-axis. Scatter plots are presented in the same order of condition importance within a phenotype from left to right. Data points represent all intervention participants in the dataset.

***Prediction Performance Metrics:*** Counts of participants who satisfy the phenotype conditions: 17 split as 17 with clinically significant response + 0 with no clinically significant response.

- precision = 17/17 (100%)
- sensitivity = 17/49 (35%)

**Responder Phenotype 1 Summary:** This phenotype predicts with 100% precision that a study participate with low back pain with the following characteristics will respond to MBSR: having low fatigue, having high self-efficacy around performing daily activities despite chronic pain (function), perceiving their general health as relatively good, having any except for the highest level of social support, and having some degree of fear and avoidance of physical activity and work.

**Figure 3:**
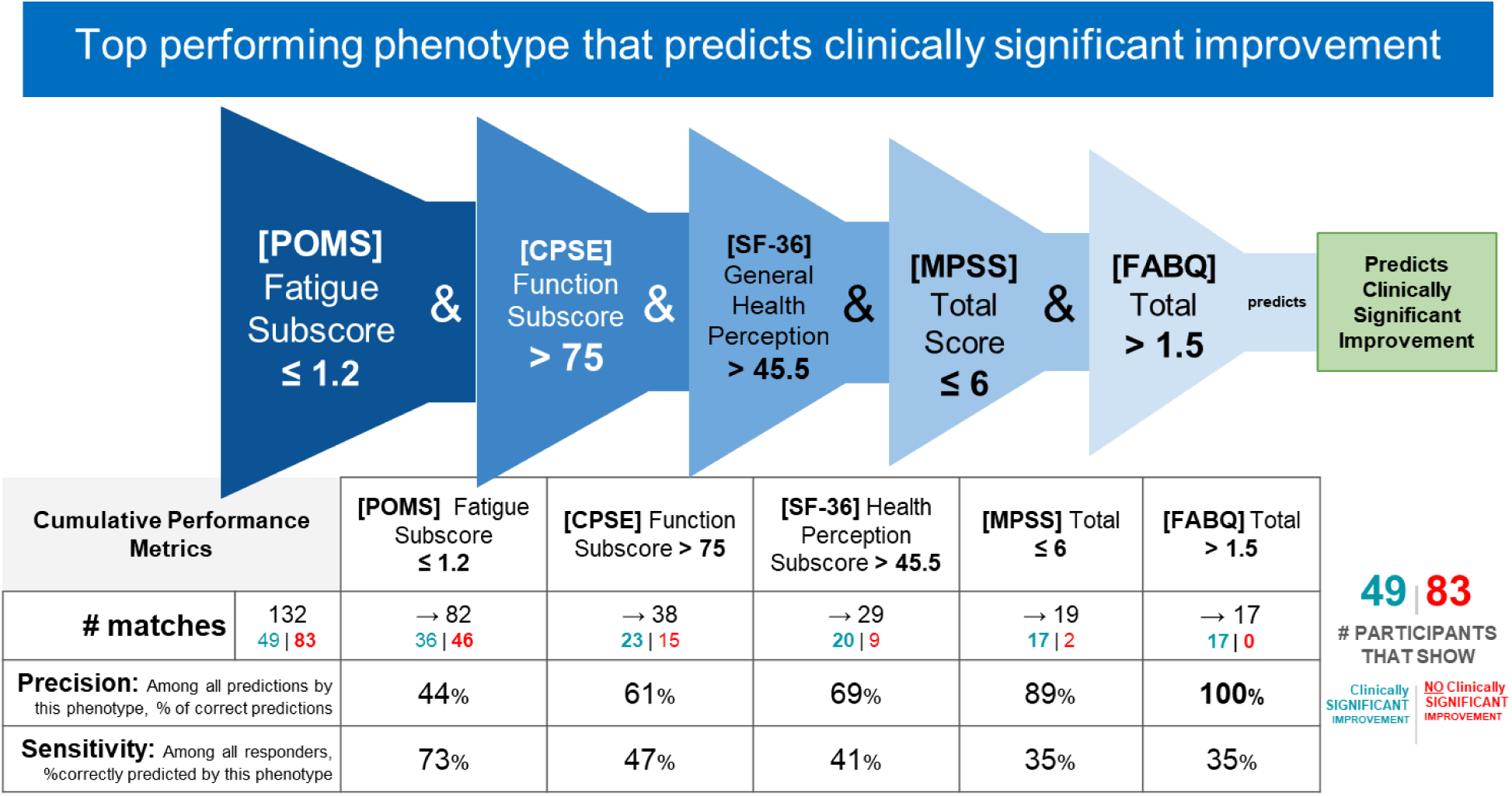
Responder Phenotype 1 Conditions and Performance Metrics. The 2nd column shows the precision and sensitivity values of the 1st phenotype condition; each subsequent column shows the precision and sensitivity values of the combined conditions from the 1st condition to that of the current column. Hence, the last column presents the total precision and sensitivity of the phenotype.

##### Responder phenotype 2 improvement in Roland Morris with MBSR

**Figure 4:**
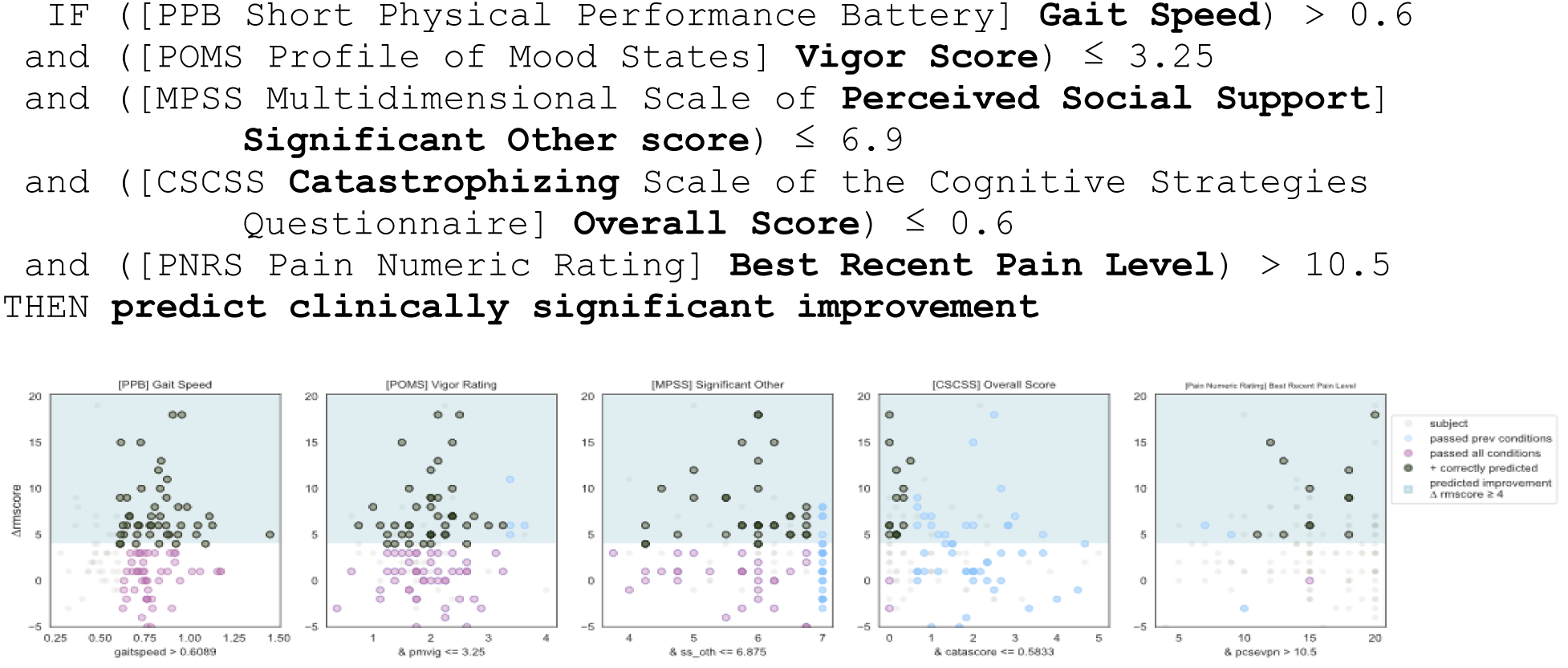
Responder Phenotype 2 Scatter Plots. Each scatter plot corresponds to one phenotype condition and depicts the phenotype condition values on the x-axis vs. the outcome (ΔRMscore) values on the y-axis. Scatter plots are presented in the same order of condition importance within a phenotype from left to right. Data points represent all intervention participants in the dataset.

***Prediction Performance Metrics:*** Counts of participants who satisfy the phenotype conditions: 13 split as 12 with clinically significant response + 1 with no clinically significant response.

- precision = 12/13 (92%)
- sensitivity = 12/49 (24%)

**Responder Phenotype 2 Summary:** This phenotype predicts with 92% precision that an individual with the following characteristics will respond to MBSR: slow to normal gait speed, having low or moderate vigor score, having any except for the highest value of social support from their significant other, extremely low to no catastrophizing, experiencing moderate or greater levels of pain.

**Figure 5:**
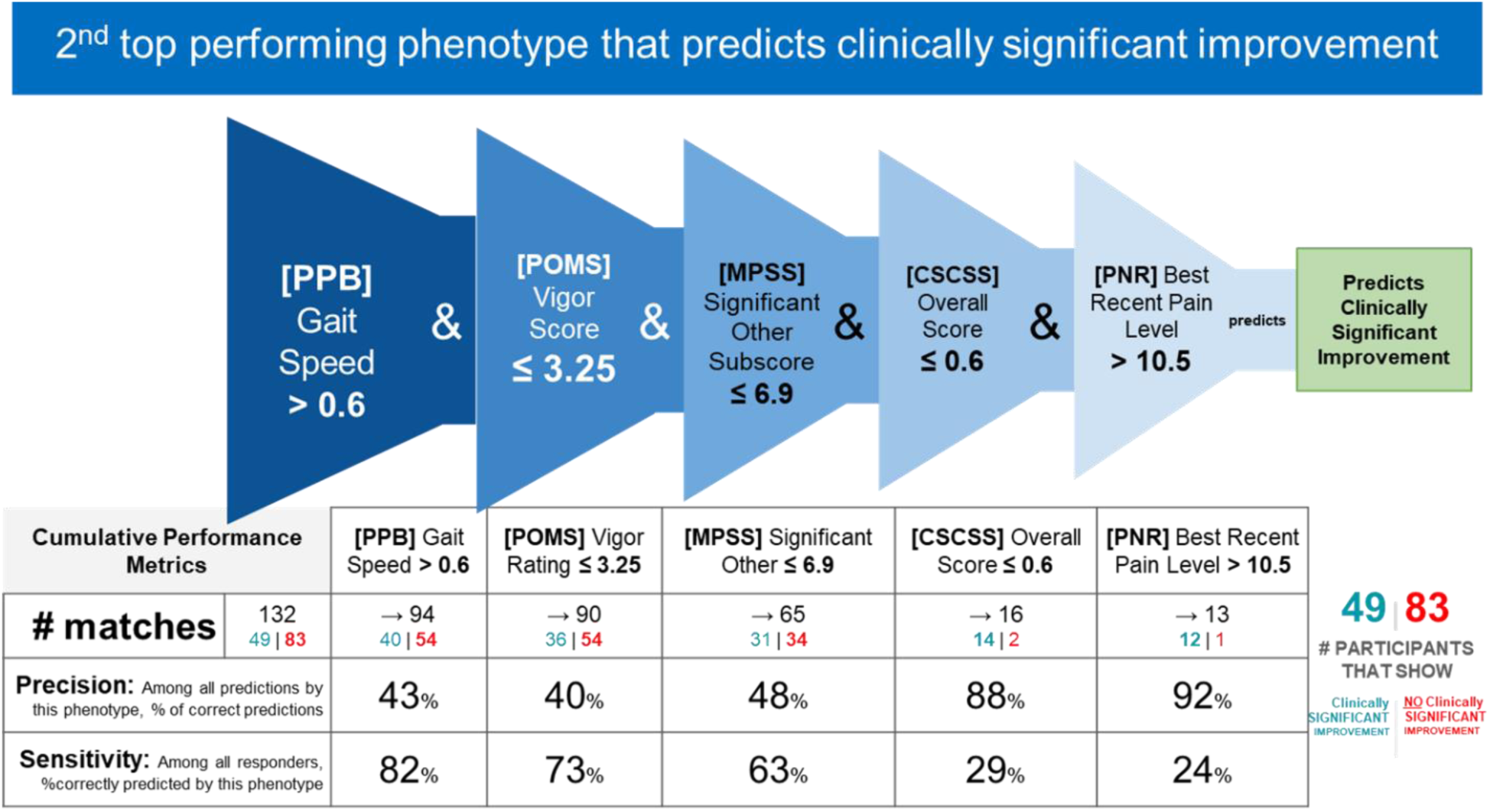
Responder Phenotype 2 Conditions and Performance Metrics. The 2nd column shows the precision and sensitivity values of the 1st phenotype condition; each subsequent column shows the precision and sensitivity values of the combined conditions from the 1st condition to that of the current column. Hence, the last column presents the total precision and sensitivity of the phenotype.

##### Responder phenotype 3 in Roland Morris with MBSR (See Figure 7)

**Figure 6:**
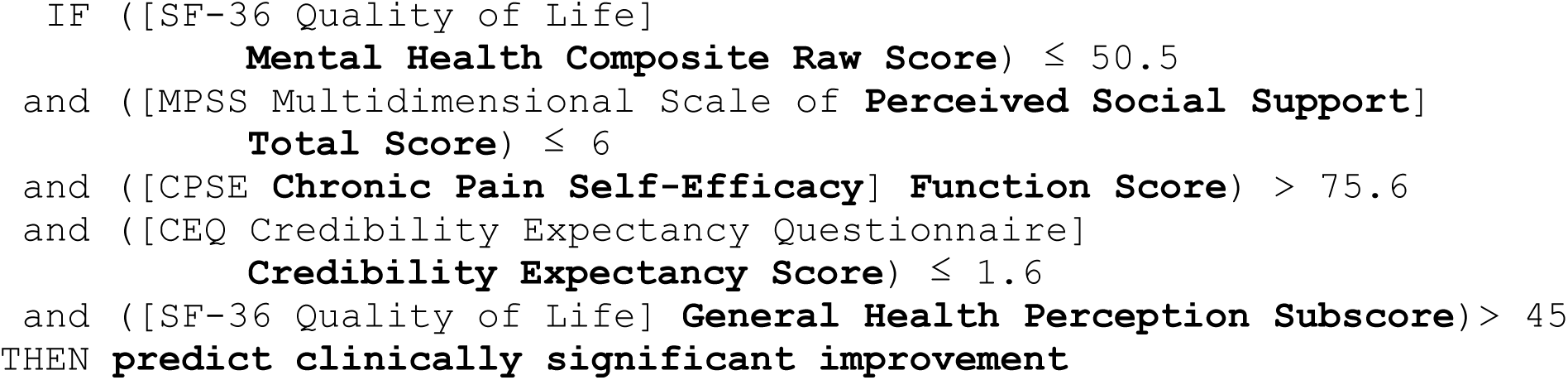
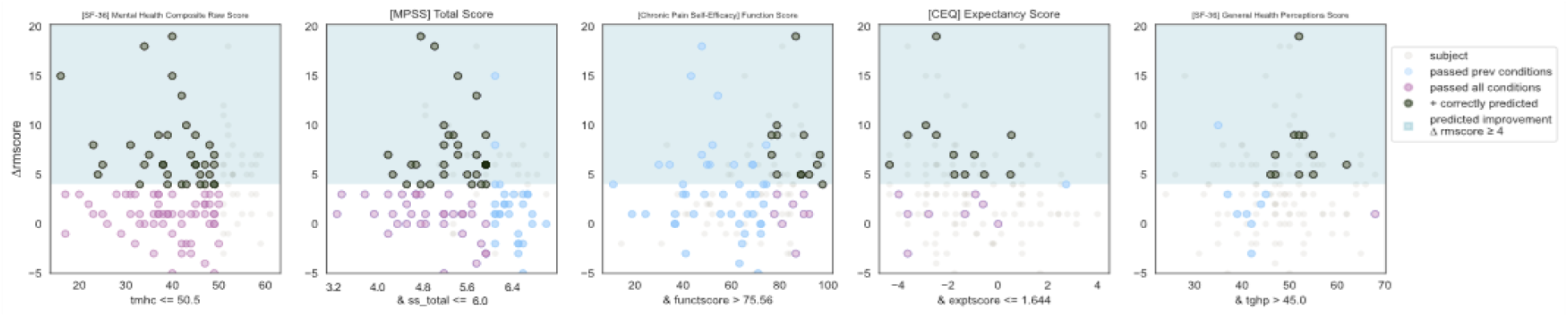
Responder Phenotype 3 Scatter Plots. Each scatter plot corresponds to one phenotype condition and depicts the phenotype condition values on the x-axis vs. the outcome (ΔRMscore) values on the y-axis. Scatter plots are presented in the same order of condition importance within a phenotype from left to right. Data points represent all intervention participants in the dataset.

***Prediction Performance Metrics:*** Counts of participants who satisfy the phenotype conditions: 12 split as 11 with clinically significant response + 1 with no clinically significant response.

- precision = 11/12 (92%)
- sensitivity = 11/49 (22%)

**Responder Phenotype 3 Summary:** This phenotype predicts with 92% precision that an individual with the following characteristics will respond to MBSR: having less than average self-perceived mental health, having any except for the highest level of social support, having high self-efficacy about the ability to function through chronic pain, having moderate to low expectancy around treatment outcomes, and perceiving their general health as relatively good.

**Figure 7:**
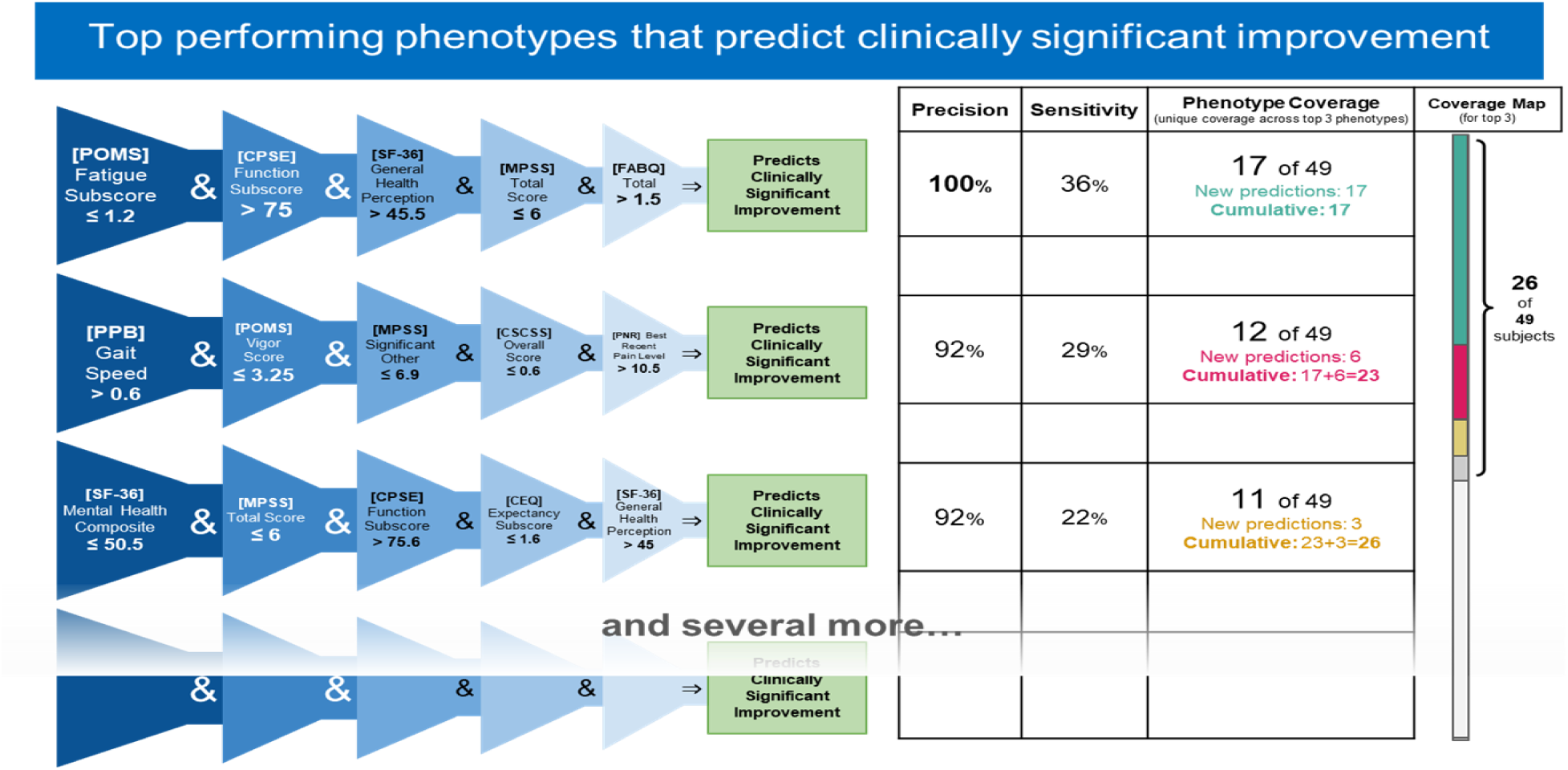
Top 3 Performing Responder Phenotypes. The rightmost column shows the coverage of each phenotype (the number of responders correctly identified by the phenotype) as well as the cumulative phenotype coverage (the number of single responders identified by the 1^st^ phenotype, by the 1^st^ and 2^nd^ phenotypes, and by the 1^st^, 2^nd^, and 3^rd^ phenotypes).

#### Top non-responder phenotypes

##### Phenotype 1 predicting no clinically significant improvement in Roland Morris with MBSR

**Figure 8:**
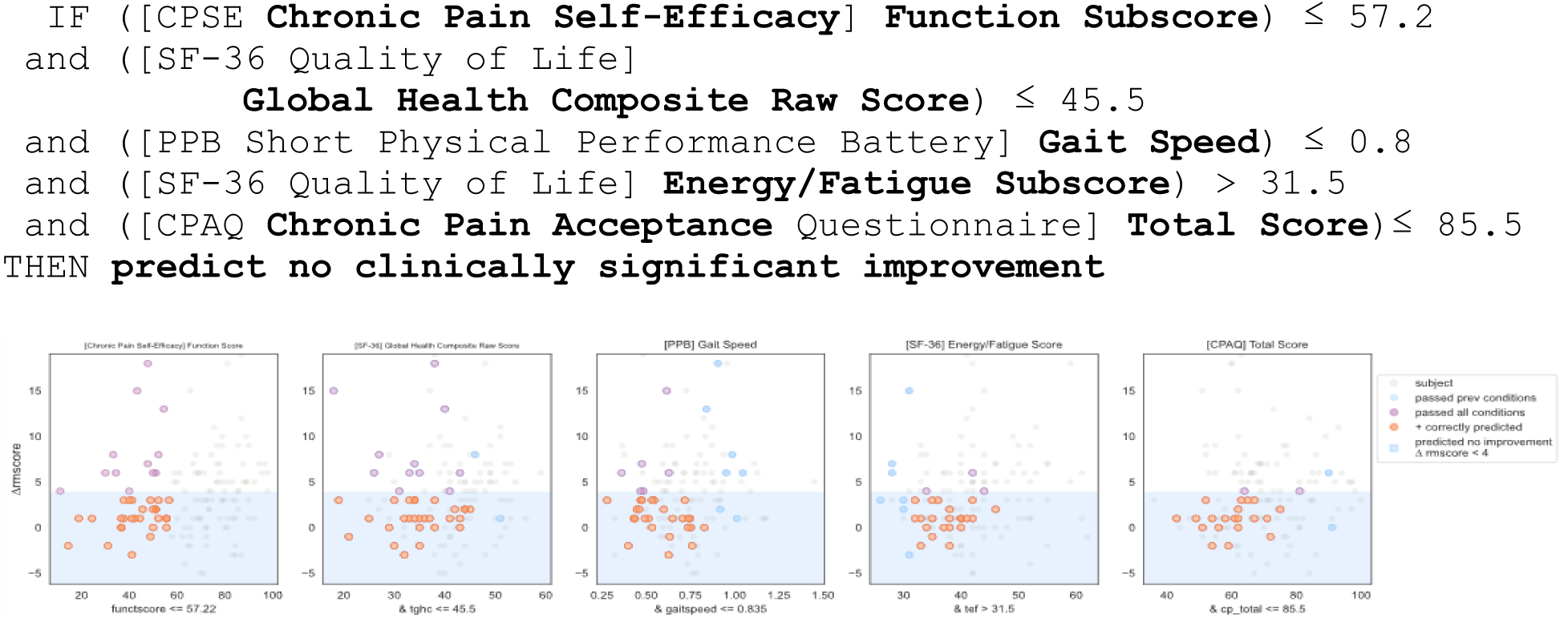
Non-responder Phenotype 1 Scatter Plots. Each scatter plot corresponds to one phenotype condition and depicts the phenotype condition values on the x-axis vs. the outcome (ΔRMscore) values on the y-axis. Scatter plots are presented in the same order of condition importance within a phenotype from left to right. Data points represent all intervention participants in the dataset.

***Prediction Performance Metrics:*** Counts of participants who satisfy the phenotype conditions: 22 split as 0 with clinically significant response + 22 with no clinically significant response.

- precision = 22/22 (100%)
- sensitivity = 22/83 (27%)

**Non-Responder Phenotype 1 Summary:** This phenotype predicts with 100% precision that an individual with the following characteristics will not response to MBSR: having low to moderate self-efficacy around performing daily activities despite chronic pain (function), low or moderate self-perception of physical and mental health, having a gait speed that limits their ambulation in the community setting, having clinically meaningful fatigue with moderate impairment, and low to moderate pain acceptance.

**Figure 9:**
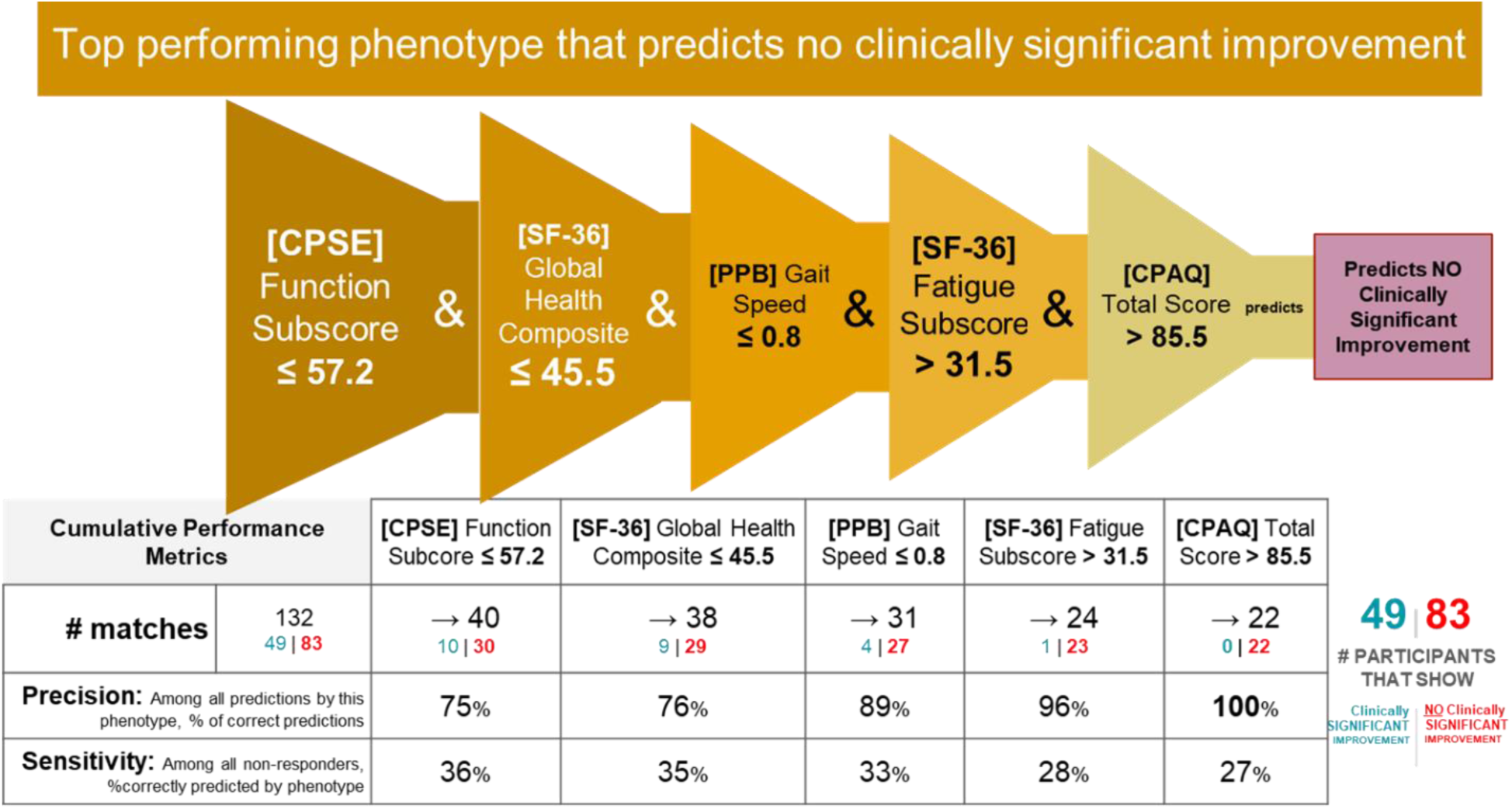
Non-responder Phenotype 1 Conditions and Performance Metrics. The 2nd column shows the precision and sensitivity values of the 1st phenotype condition; each subsequent column shows the precision and sensitivity values of the combined conditions from the 1st condition to that of the current column. Hence, the last table column presents the total precision and sensitivity of the phenotype.

##### Phenotype 2 predicting no clinically significant improvement in Roland Morris with MBSR

**Figure 10:**
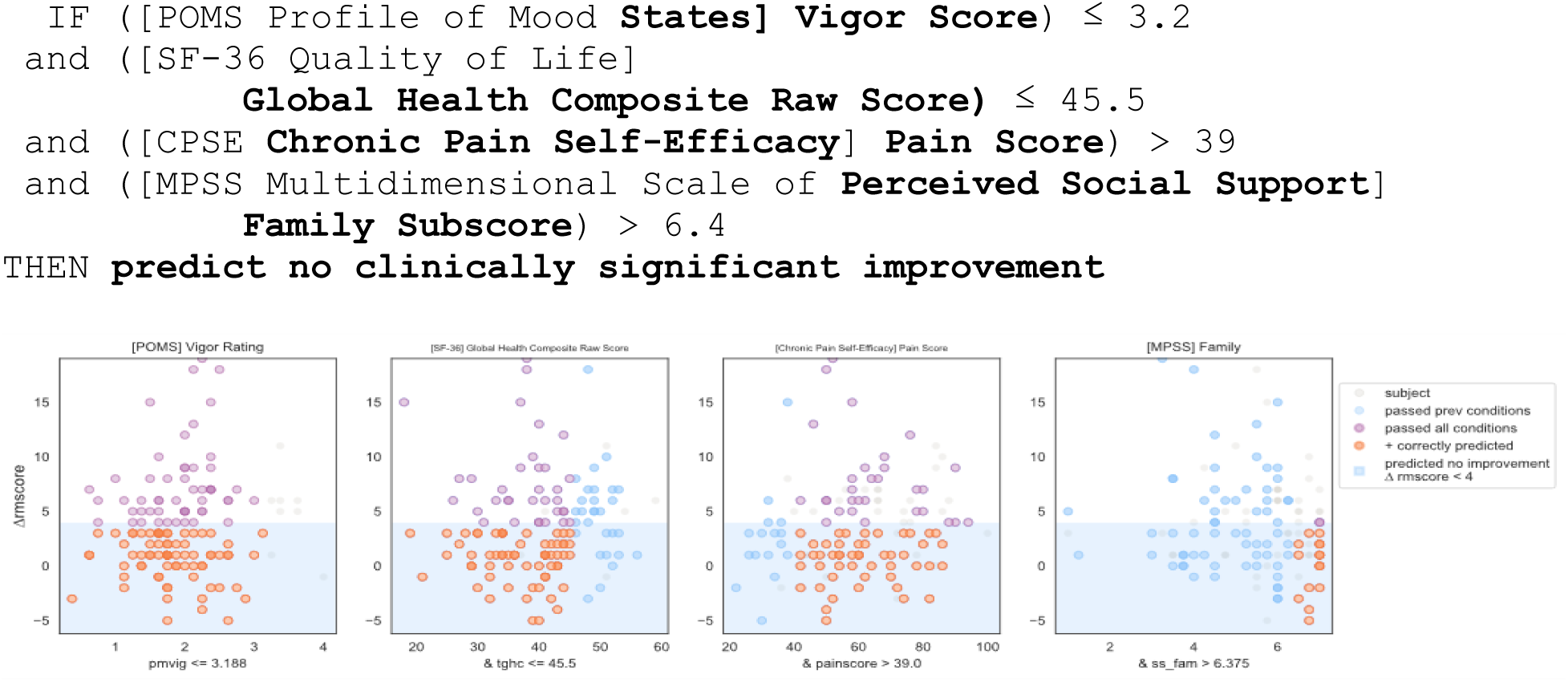
Non-responder Phenotype 2 Scatter Plots. Each scatter plot corresponds to one phenotype condition and depicts the phenotype condition values on the x-axis vs. the outcome (ΔRMscore) values on the y-axis. Scatter plots are presented in the same order of condition importance within a phenotype from left to right. Data points represent all intervention participants in the dataset.

***Prediction Performance Metrics:*** Counts of participants who satisfy the phenotype conditions: 17 split as 0 with clinically significant response + 17 with no clinically significant response.

- precision = 17/17 (100%)
- sensitivity = 17/83 (20%)

**Non-Response Phenotype 2 Summary:** This phenotype predicts with 100% precision that an individual with the following characteristics will not response to MBSR: having low or moderate vigor, having average or lower physical and mental health, having moderate or high self-efficacy around living with chronic pain, and having a high degree of social support from family.

**Figure 11:**
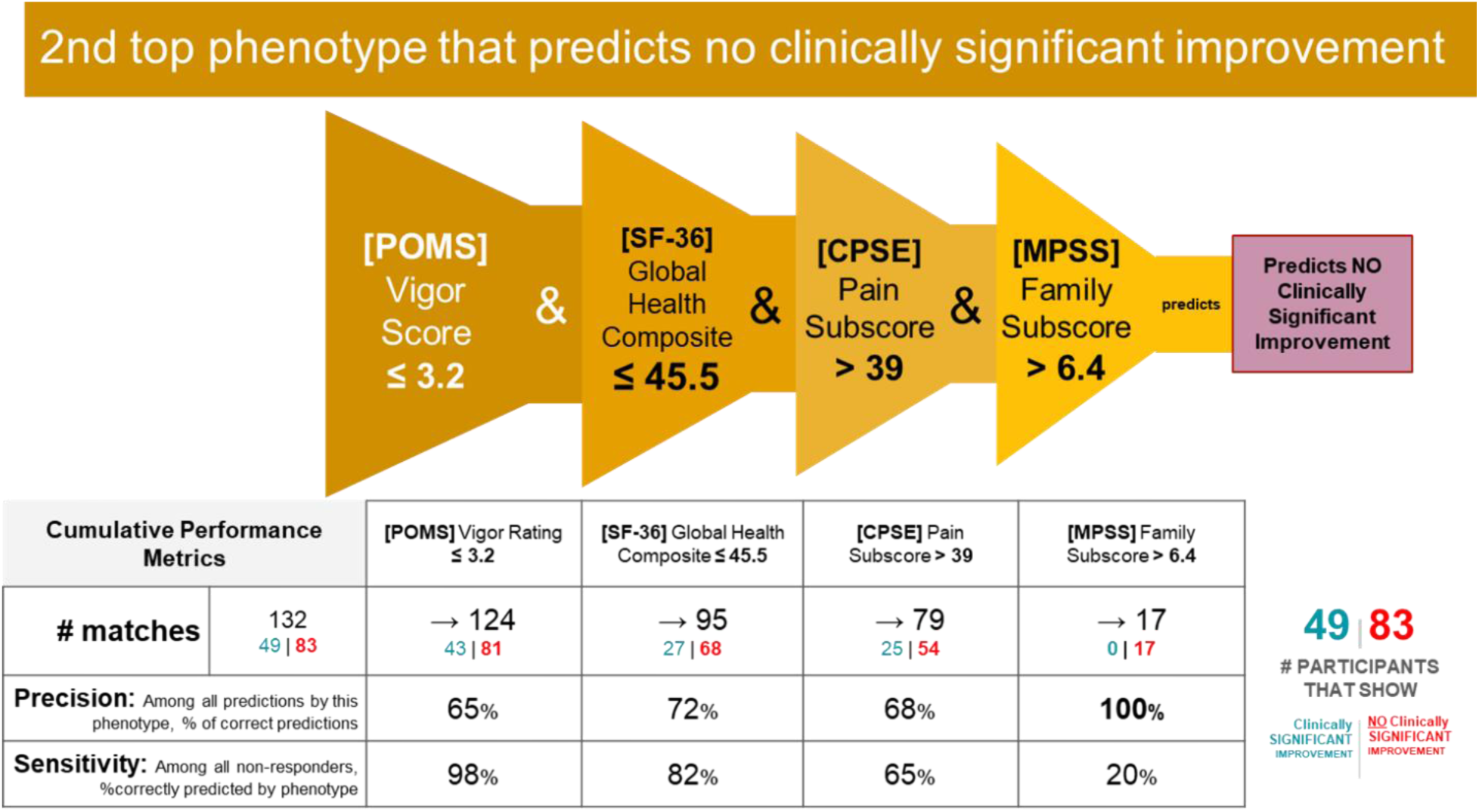
Non-responder Phenotype 2 Conditions and Performance Metrics. The 2nd column shows the precision and sensitivity values of the 1st phenotype condition; each subsequent column shows the precision and sensitivity values of the combined conditions from the 1st condition to that of the current column. Hence, the last column presents the total precision and sensitivity of the phenotype.

##### Phenotype 3 predicting not clinically significant improvement in Roland Morris with MBSR (see figure 13)

**Figure 12:**
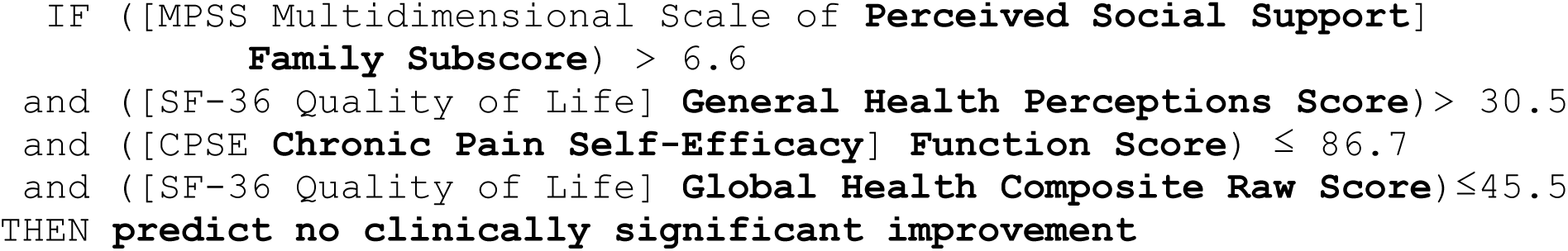
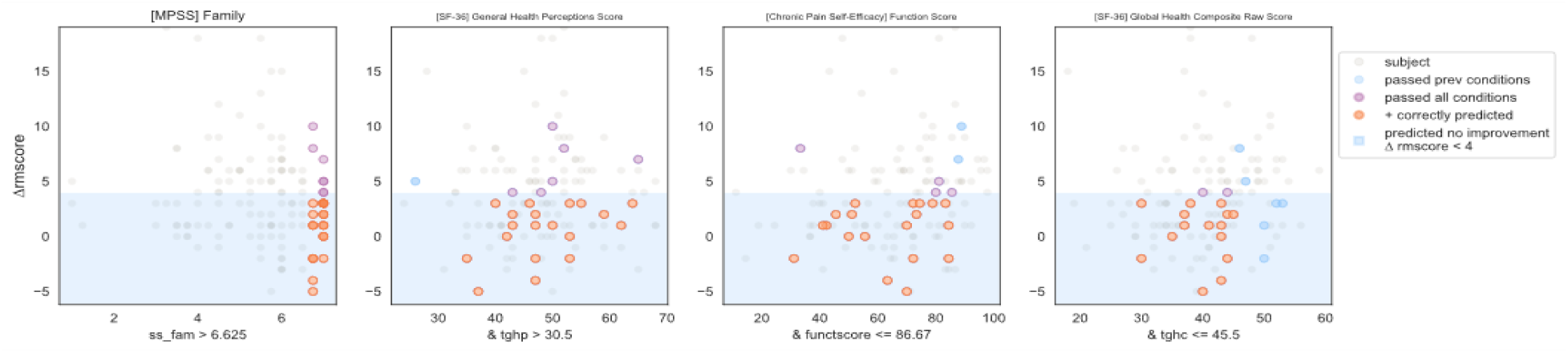
Non-responder Phenotype 1 Scatter Plots. Each scatter plot corresponds to one phenotype condition and depicts the phenotype condition values on the x-axis vs. the outcome (ΔRMscore) values on the y-axis. Scatter plots are presented in the same order of condition importance within a phenotype from left to right. Data points represent all intervention participants in the dataset.

***Prediction Performance Metrics:*** Counts of participants who satisfy the phenotype conditions: 17 split as 0 with clinically significant response + 17 with no clinically significant response.

- precision = 17/17 (100%)
- sensitivity = 17/83 (20%)

**Figure 13:**
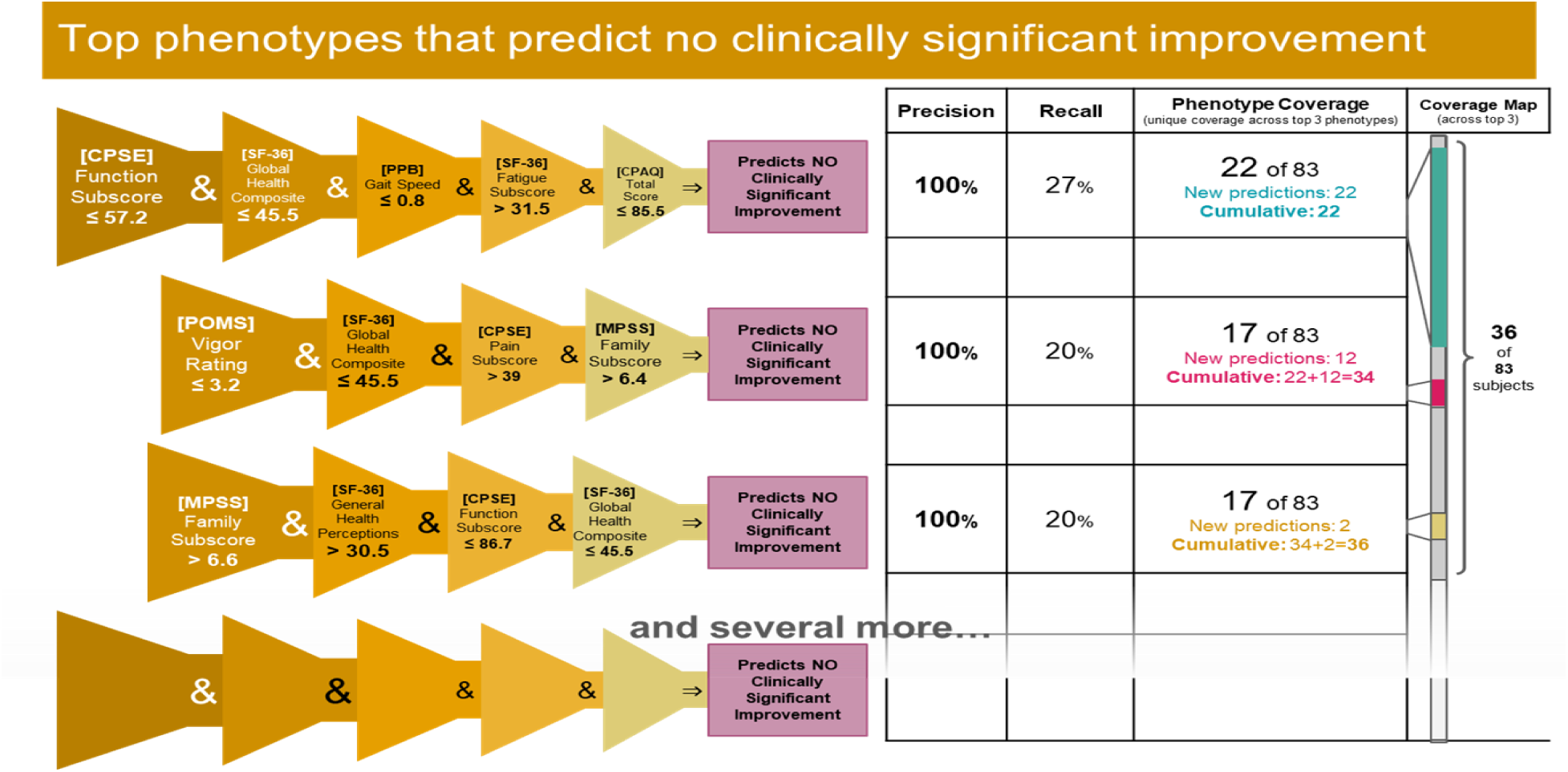
Top 3 Performing Non-responder Phenotypes. The rightmost column shows the coverage of each phenotype (the number of non-responders correctly identified by the phenotype) as well as the cumulative phenotype coverage (the number of single non-responders identified by the 1^st^ phenotype, by the 1^st^ and 2^nd^ phenotypes, and by the 1^st^, 2^nd^, and 3^rd^ phenotypes)

**Non-Response Phenotype 3 Summary:** This phenotype predicts with 100% precision that an individual with the following characteristics will not response to MBSR: having a high degree of social support from family, self-perceiving their general health as moderate or high, having a wide range except for the highest level of self-efficacy around functioning with pain, and having low or moderate perception of physical and mental health.

## DISCUSSION

### Overall Summary and Performance

The present analyses demonstrate the utility of machine learning approaches for accurately identifying responders and non-responders to mindfulness for cLBP. This was achieved through constructing predictive phenotypes encompassing clusters of relevant characteristics. Three responder phenotypes covered over 50% of the responders with 92-100% precision, and three non-responder phenotypes covered 43% of non-responders with 100% precision. This required only four-to-five variables, where precision was still 80-100% with four variables. Our results can be used to inform future related studies and patient and clinician decision making on mindfulness interventions for chronic pain to maximize clinical efficacy, patient outcomes, and resource use.

### Contributions to predictive ML studies of chronic pain

While there is robust literature on the use of ML to predict pain levels in general, studies predicting the response to interventions for pain reduction, especially complementary interventions, are limited. Furthermore, most investigations focus on identifying individual or groups of predictive factors rather than specific phenotypes. Phenotypes add granularity around not only specific clusters of features, but also thresholds above or below which a particular feature becomes meaningful. The current phenotype development therefore provides enhancements in both specificity and accuracy compared to many earlier studies, increasing reliability and confidence in related clinician and/or patient decision making. In clinical settings, if a patient closely matches a certain highly-predictive phenotype, this could be a strong indicator that mindfulness is a favorable option for that individual. Similarly, if a patient matches multiple phenotypes, this would additionally increase confidence in likelihood of response. The presented methods can also be easily applied to other populations and/or types of interventions to derive similar specificity and accuracy and integrated into automated algorithms to guide decision-making.

### Responder phenotypes

Common responder attributes found in multiple phenotypes consisted of high chronic pain self-efficacy, moderate to high perceptions of overall health, any except for the highest level of social support, and low treatment expectancy. Other attributes included low fatigue and catastrophizing, low to moderate perceived mental health, and moderate or greater recent pain level. Taken together, this suggests that individuals who respond to mindfulness are, at baseline, more confident in their ability to manage their own pain, perceive themselves to be in good health, have some social support, and do not have high expectations of the mindfulness intervention.

The high self-efficacy and perception of overall health may indicate that responders have the motivation and optimism necessary for a behavioral intervention such as mindfulness to work (Wright and Schutte, 2014). Although treatment expectancy plays a significant role in how individuals respond to mindfulness-based interventions (MBIs) for pain relief, where positive expectations about the effectiveness of mindfulness can contribute to greater pain reduction (Peerdeman et al., 2016, Lopes et al., 2024), the lack of high expectations in the present responder phenotypes may make it less likely they will be disappointed in the treatment outcome. In some phenotypes, this is associated with low fatigue and catastrophizing, not surprising given the internal motivation and optimism necessary for mindfulness-based interventions to have significant effects (Finan and Garland, 2015). Furthermore, the relationship between mindfulness and back pain-associated disability may be mediated by catastrophizing (Cassidy et al., 2012).

An enhanced ability to identify responders to mindfulness interventions for cLBP has clear benefits to patients and providers, including accelerated rate of symptom improvement and lower chances of progression to other higher risk interventions, such as surgery and/or opioids. The communication and discussion of suitability for mindfulness interventions between clinician and patient may increase expectancy, improving outcomes. A general increased awareness of predictors of a positive response to mindfulness interventions among providers may broaden the application of these effective, low-risk treatments.

### Non-responder phenotypes

Common non-responder attributes identified in multiple phenotypes consisted of moderate to low chronic pain self-efficacy, moderate to low perception of overall health, and the highest level of social support from family. Other attributes included slow gait, low vigor, and moderate or lower chronic pain acceptance. The low pain self-efficacy may be indicative of an inability to take advantage of beneficial aspects of mindfulness related to improved self-care (Zeidan and Vago, 2016, Sawyer, 2023). Lower perception of overall health and low pain acceptance in the non-responder phenotypes could indicate general health related pessimism and inability to benefit from the acceptance elements of mindfulness for chronic pain (Ploesser and Martin, 2024, Kober et al., 2020). Self-efficacy and pain acceptance are associated with mindfulness attributes, and self-efficacy may mediate the relationship between mindfulness and emotion regulation (Turner et al., 2016). The common presence of social support from family in the non-responder phenotypes could indicate that individuals with social support from relatives (possibly beyond that of a significant other) may not benefit from social support aspects of mindfulness interventions.

The ability to accurately identify non-responders to mindfulness for cLBP presents multiple distinct benefits. In addition to directing treatment away from those least likely to benefit from these often time-and resource-intensive interventions, it can be used to identify non-responder populations and to inform adjunct interventions or tailor the mindfulness programming to potentially address the causes, moderators, and/or mediators of non-responsiveness. In this vein, if a patient closely matches one or more non-response phenotypes, then the clinical goal might shift toward first addressing any modifiable factors that predict non-response to improve the patient’s suitability for mindfulness treatment. For example, if it is determined that low pain self-efficacy inhibits a positive response to mindfulness, the intervention could be modified to increase the emphasis on improving self-efficacy, either before or in parallel with the mindfulness.

### Strengths, Limitations, and Future Work

These results make a novel contribution to the field and forward the goal of achieving a personalized and precise understanding of response to mindfulness for chronic pain. Nonetheless, while the present analyses successfully and accurately identified predictive phenotypes with high precision and good sensitivity, the intervention sample was still modest in size and comprised of older adults. Further comparative and validation work is needed to determine how well these and similar phenotypes can predict response or non-response in other relevant at-risk clinical populations. An increased emphasis on the trajectory of predictors (change over time in addition to baseline values) would provide further insight into how to address the lack of response to mindfulness for chronic pain, including through identifying potential pre-intervention targets for intervention. That is, modifying certain features that are predictive of non-response may increase someone’s likelihood that mindfulness would subsequently be effective for reducing pain. We also did not have access to various other types of data, such as physical activity, neural activity or blood-based biomarkers, which could improve precision and/or sensitivity. Ongoing and future work should capitalize on other data modalities including activity and sleep sensors, omics, and neuroimaging as well as train models with larger clinical datasets. These additions would augment the robustness of machine learning approaches for identifying useful clusters of predictive characteristics.

### Conclusions

Results from this machine learning based phenotyping provide an integrated portrait of responders and non-responders. These phenotypes – and those that may emerge from using machine learning in other datasets, can meaningfully inform clinician and patient decision making to ultimately promote better pain outcomes for patients with chronic pain. This work can also more broadly inform research and development of mindfulness-based treatments for pain and advance the central goal of personalized, complementary pain management.

## Data Availability

All data produced in the present are available upon reasonable request to the authors.

## FIG

